# Improving Motivation in Post-stroke Apathy with Repetitive Transcranial Magnetic Stimulation (IMPART): A Phase-I Pilot Trial

**DOI:** 10.64898/2026.06.01.26354398

**Authors:** Michael B. Seidman, Parneet Grewal, Colin Bowyer, Ivy Dickens, Jacob Eade, Elisabeth Collins, Cina Patel, Diego Eduardo Arias Velasquez, Mark S. George, Michael Antonucci, Kevin A. Caulfield, Lisa M. McTeague

**Affiliations:** Department of Neurology, University of Colorado, Aurora, Colorado; Department of Neurology, Medical University of South Carolina, Charleston, SC; Department of Psychiatry & Behavioral Sciences, Medical University of South Carolina, Charleston, SC; College of Medicine, Medical University of South Carolina, Charleston, SC; College of Health Professions, Medical University of South Carolina, Charleston, SC; Research Service, Ralph H. Johnson VA HCS, Charleston, SC; Department of Neuroradiology, Medical University of South Carolina, Charleston, SC

**Keywords:** cerebrovascular disease, stroke, apathy, transcranial magnetic stimulation, rTMS, motivation, chronic stroke, intermittent theta burst

## Abstract

**Background:** Post-stroke apathy (PSA) is a common, disabling syndrome with few evidence-based treatment options. We evaluated the safety, feasibility, acceptability, and evidence of effects of a three-day accelerated intermittent theta burst stimulation-repetitive transcranial magnetic stimulation (iTBS-rTMS) protocol targeting the left dorsomedial prefrontal cortex (dmPFC) in chronic stroke survivors with apathy.

**Methods:** Stroke survivors with symptomatic apathy received open-label iTBS-rTMS at the left dmPFC (21,600 pulses across 36 sessions; 3 treatment days; 12 sessions/day within one week). Safety endpoints included adverse events, neuroradiological findings, and objective cognitive performance. Secondary outcomes included measures of apathy and other neuropsychiatric symptoms, as well as psychosocial functioning, including quality of life and caregiver burden. Participants were followed up for one month.

**Results:** Fourteen participants (mean age = 61.8 ± 14.0 years; mean time since stroke = 55.6 ± 31.6 months) completed the iTBS-rTMS treatment course. No serious adverse events occurred. Participants rated the treatment as highly acceptable, and cognitive performance was stable from pre- to post-rTMS, with no treatment-related changes on structural MRI. Regarding apathy, participants had significant improvements with moderate to large effect sizes on the Lille Apathy Rating Scale (LARS), on both self (*d* = 0.78) and caregiver-rated versions (*d* = 1.28), p<0.05 pretreatment-to-one-month follow-up. In addition, secondary measures of psychosocial function also showed improvement with moderate to large effect sizes (Stroke Specific Quality of Life Scale: *d* = 0.62; Zarit Burden Interview: *d* = 0.72), and the Brief Inventory of Psychosocial Function: *d* = 0.89).

**Conclusions:** In chronic stroke survivors with PSA, accelerated iTBS-rTMS targeting the left dmPFC appears to be safe, feasible, tolerable, and highly acceptable, with preliminary evidence suggesting a potential role in reducing apathy and secondarily promoting improvements in quality of life, caregiver burden, and broader psychosocial function.

Clinical Trial Registration ID: NCT05878457, listing: https://clinicaltrials.gov/study/NCT05878457

**Nonstandard Abbreviations and Acronyms:** none

**Clinical Perspective:** *What Is New?:*

- Dorsomedial prefrontal cortex (dmPFC) holds promise as a network-informed target capable of stimulation with transcranial magnetic stimulation for reducing apathy.
- High dose of intermittent theta burst (iTBS) stimulation administered to date in a chronic stroke population to dmPFC appears safe, well-tolerated, and feasible with a novel accelerated protocol.

*What Are the Clinical Implications?:*

- Accelerated iTBS at the left dmPFC appears to be a feasible and potentially efficacious target for apathy in chronic stroke—a patient group with limited evidence-based intervention options.

## INTRODUCTION

Apathy is a common, disabling neuropsychiatric syndrome common in stroke, characterized by reduced goal-directed behaviors across emotional, cognitive, behavioral, and/or social domains.^1,2^ Post-stroke apathy (PSA) affects over one-third of stroke survivors within five years, and is independently associated with a host of negative outcomes, including diminished functional recovery, rehabilitation engagement, and quality of life, coupled with increased morbidity and mortality.^3–9^ There are currently no FDA-approved treatments for PSA, and existing off-label pharmacologic strategies show mixed evidence^10^ and risk undesirable side effects. Non-pharmacological interventions such as rehabilitative therapies show promise, but are likewise limited by incomplete evidence,^10^ and may be time-consuming or difficult to access for stroke survivors, who are often restricted by physical disability and limited transportation.^11,12^

A growing body of evidence suggests that the neuroanatomical substrates of apathy converge on frontal-subcortical circuits subserving goal-directed behavior, with the anterior cingulate cortex (ACC) serving as an integratory hub.^13–16^ The disruption of this motivational network at various nodes and even via downstream inputs from distant effectors is thought to lead to common apathy phenotypes across diverse clinical populations, and may explain the ubiquity of apathy among stroke survivors despite heterogeneous lesion locations.^15–17^

Repetitive transcranial magnetic stimulation (rTMS) is a non-invasive neuromodulation technique that can stimulate cortical targets of interest to induce downstream changes in network-level functional connectivity and synaptic plasticity.^18,19^ While a traditional course of rTMS consists of daily sessions delivered over a period of 6-7 weeks, recent innovations, including intermittent theta burst stimulation (iTBS) and accelerated protocols, allow for the delivery of an equivalent course over a period of days rather than weeks, significantly reducing treatment burden with comparable safety and tolerability.^20^ rTMS is well-established as safe and tolerable in primary psychiatric populations, with a growing body of evidence supporting numerous primary neurologic indications as well, including stroke and apathy specifically.^21–25^

Among cortical regions accessible to stimulation via rTMS, the dorsomedial prefrontal cortex (dmPFC) may represent a particularly appealing target for treating apathy transdiagnostically, given its interconnectedness with the ACC centrally within the motivational network. This is especially important in stroke, as heterogeneous lesions may compromise the motivational network through variable pathways.^17^ A signal for dmPFC-targeted rTMS as a potentially feasible and efficacious treatment for PSA comes from a 2017 study by Sasaki and colleagues,^22^ but accelerated, high-dose iTBS at the dmPFC has never been investigated in a chronic stroke population or for PSA specifically. We therefore conducted an open-label phase I trial to investigate whether accelerated rTMS at the left dmPFC is safe and feasible, and to provide early evidence of its potential to remediating apathy in chronic stroke.

## METHODS

### Data Availability

Anonymized data not published within this article will be available from the corresponding author upon reasonable request from any qualified investigator.

### Study Design

This was an open-label phase I pilot study designed to evaluate the safety, feasibility, tolerability, acceptability, and preliminary efficacy of accelerated iTBS-rTMS at the left dmPFC for PSA (**Figure 1A**). The study was conducted at the Medical University of South Carolina (MUSC), a comprehensive stroke center in South Carolina, and was approved by the MUSC Institutional Review Board (IRB; Pro00126436). The study adhered to the Declaration of Helsinki and Consolidated Standards of Reporting Trials (CONSORT) extension for pilot and feasibility trials and was registered on ClinicalTrials.gov (NCT05878457).

**Figure 1:**
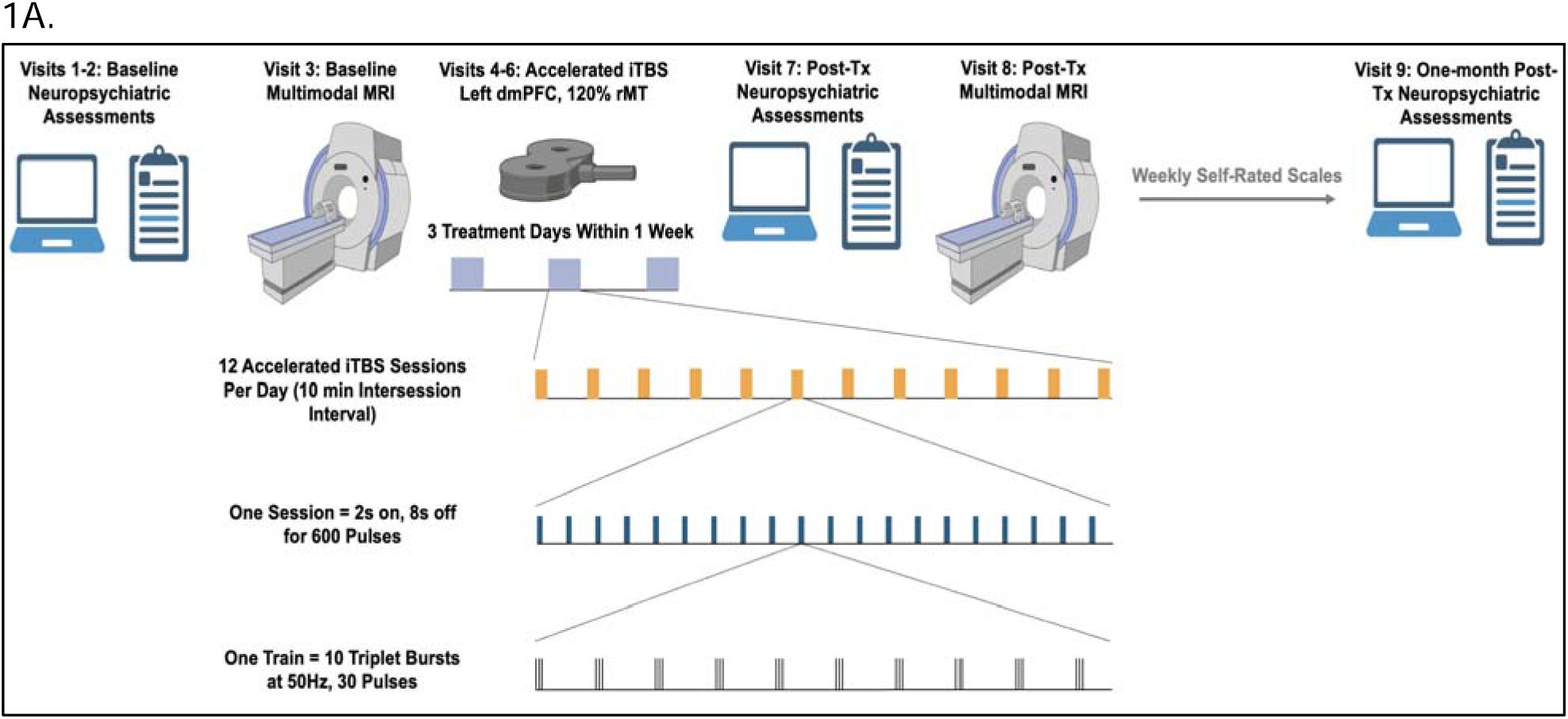
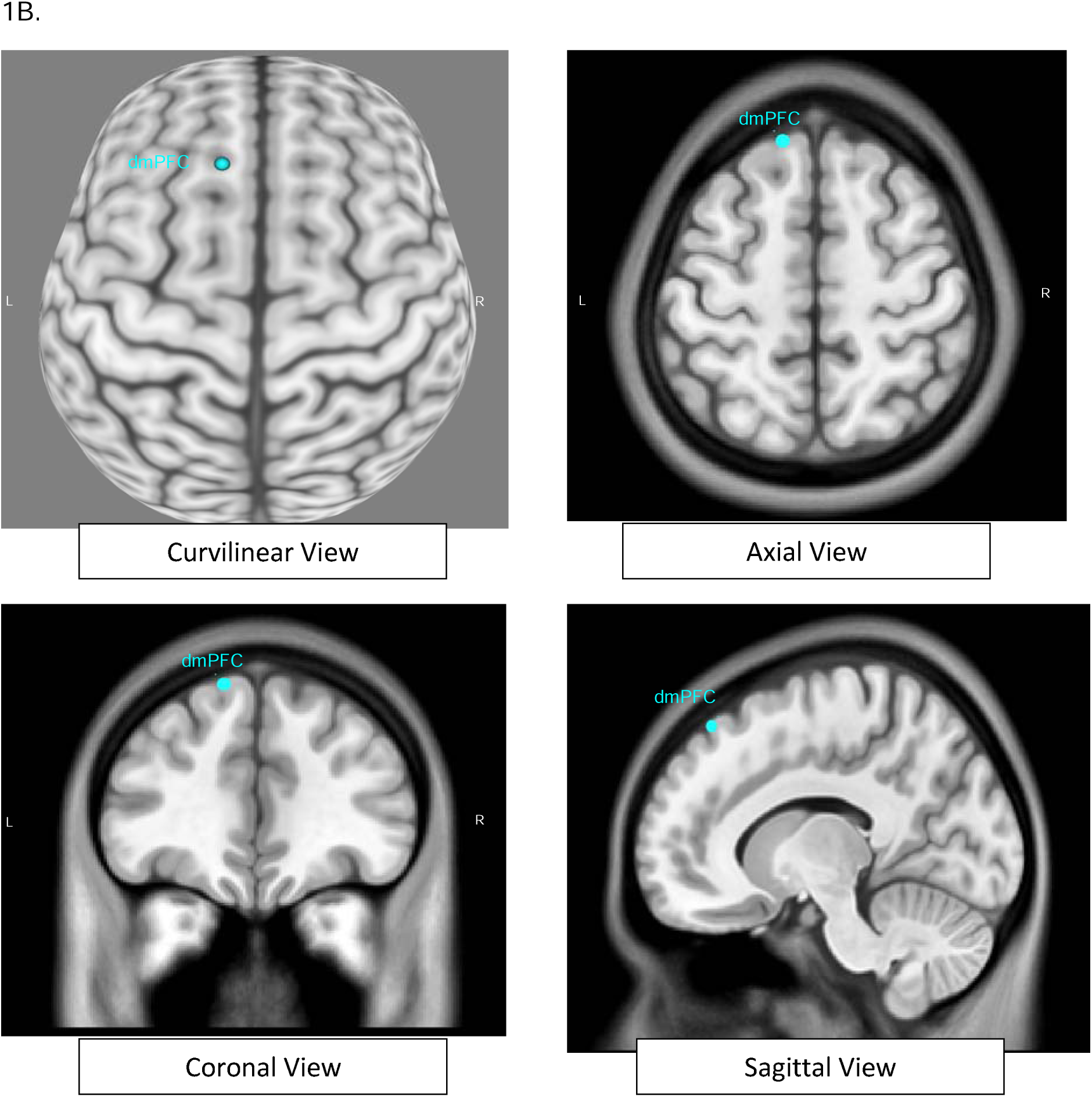
Study design and procedures. **1A**. Study timeline including pre-screening, baseline assessments, and follow-up. **1B**. Neuronavigation using the Brainsight Neuronavigation system (Version 2.4.8; Rogue Research, Montreal, QC, Canada) showing the location of Dorsomedial Prefrontal Cortex (dmPFC) with MNI coordinates (curvilinear, axial, coronal, and sagittal views).

### Participant Eligibility

Participants were enrolled with a co-participant informant (e.g., care partner, family member, or close friend) who completed collateral assessments throughout the study. Inclusion criteria for primary participants were: age ≥ 40 years; ischemic or hemorrhagic stroke with ≥ 6 months chronicity; clinically significant apathy (Apathy Evaluation Scale [AES] ≥ 39 on self- or informant-rated forms^26^); ability to complete psychometric testing; and an intact cortex at the site of stimulation confirmed by pre-treatment Magnetic Resonance Imaging (MRI). Exclusion criteria included primary extra-axial hemorrhage without evidence of intracerebral ischemic or hemorrhagic stroke; other concomitant neurological diagnoses significantly affecting cognitive or motor function; moderate-to-severe global aphasia; visual impairment precluding cognitive testing; contraindications to MRI or rTMS; pregnancy; seizure disorder; scalp lesions, bone defects, wounds, or hemicraniectomy at the stimulation site; claustrophobia precluding MRI; active substance use disorder; psychotic disorder; bipolar I disorder; or acute suicidality. Home medications were reviewed prior to inclusion in the trial, but no changes to home medications were made during the trial. Co-participants were required to be ≥ 18 years of age, have at least weekly in-person contact with the primary participant, and be able to provide reliable collateral assessments.

### Recruitment and Consent

Participants were recruited from the Registry for Stroke Recovery, an institutional database of stroke survivors interested in research, as well as through medical record review, clinician referral, and community outreach. Potential participants and co-participants were screened by telephone for preliminary eligibility and enrolled after providing written informed consent.

### Study Procedures

Following phone screening and consent, participants completed a battery of baseline clinical assessments, including evaluations of apathy, psychiatric symptoms, psychosocial function, quality of life, and cognitive function, followed by a baseline MRI. Open-label accelerated iTBS-rTMS was delivered on three treatment days scheduled flexibly within a 7-day treatment window. A post-treatment repeat MRI was obtained within 3 days of completion of treatment sessions. The clinical assessment battery was repeated immediately post-treatment and at 1-month post-treatment. Participants and co-participants also completed weekly self-reported apathy questionnaires during the 1-month follow-up window. Study procedures were performed by the research assistant certified under the guidance of the principal investigator and by the co-investigators, who were all certified in delivering transcranial magnetic stimulation. **(Figure 1A)**

### Accelerated iTBS-rTMS Parameters

Participants received open-label iTBS with a MagVenture MagPro® X100 Magnetic Stimulator equipped with a liquid-cooled Cool B65 figure-8 coil. Stimulation targeted the left dmPFC using a Brainsight Neuronavigation system at Montreal Neurological Institute (MNI) coordinates approximating the left dmPFC, which was approximated as MNI coordinates correlated with the midpoint between the F1 and Fz positions of the international 10-10 EEG system based on extant literature (**Figure 1B**).^27^ Treatment consisted of 21,600 total pulses delivered across 36 sessions, with 12 sessions per day on each of three consecutive or non-consecutive treatment days within a 7-day window. Each session consisted of 600 iTBS pulses, delivered as 20 trains of 30 pulses in 50 Hz triplet bursts every 200 ms (5 Hz theta frequency) with 2-second trains and 8-second inter-train intervals for 190 seconds. Sessions were separated by 10-15-minute inter-session intervals. The target stimulation intensity was fixed at 120% of the resting motor threshold (rMT). Stimulation intensity was started at either the target or a lower level (between 100-120% rMT) and ramped up gradually according to patient comfort and preference. All participants reached 120% rMT by the end of the treatment.

#### Sample Characterization

All participants completed a demographics questionnaire. Baseline cognitive function was assessed using the National Institute of Neurological Disorders and Stroke – Canadian Stroke Network (NINDS-CSN) 30-minute neuropsychological battery. Pre-stroke apathy and dementia were screened using a modified form of the Short Form of the Informant Questionnaire on Cognitive Decline in the Elderly (IQCODE). Premorbid psychiatric conditions were screened using the Quick Structured Clinical Interview for DSM-5 Disorders (SCID).

#### Primary outcome measures

##### Safety

Adverse events were recorded. Each participant underwent a non-contrast structural MRI pre- and post-treatment on a 3T Prisma^fit^ MRI system with 32-channel head coil (Siemens Healthineers, Erlangen, Germany), including volumetric T1-weighted, T2-weighted, fluid-attenuated inversion recovery, diffusion-weighted, and susceptibility-weighted images to identify any pre- to post-treatment changes or incidental findings. Electric field (E-field) modeling was performed to estimate the distribution and magnitude of the stimulation and to confirm that the induced fields were within established safety limits. Emergence of suicidality or mania was monitored with the Columbia Suicide Severity Rating Scale (C-SSRS) and the Young Mania Rating Scale (YMRS), respectively. Cognitive function was monitored using the NIH Toolbox Cognition Battery, the Montreal Cognitive Assessment (MoCA), and the Patient-Reported Outcome Measurement Information System (PROMIS) short-form cognition scale.

##### Feasibility

Feasibility was defined as successful recruitment and a retention rate of >80% among participants allocated to treatment.

##### Tolerability

A Momentary Assessment of rTMS was administered after every session on each treatment day, in which participants rated common rTMS side effects, including headache, pain, and fatigue, that they experienced both during and after rTMS. Additionally, a general review of systems was administered at the beginning of each treatment day to monitor for existing and emergent physical symptoms.

##### Acceptability

A TMS Experience Questionnaire was completed immediately post-treatment, in which participants retrospectively rated their perceptions of the treatment, including perception of side effects, benefits, ease of undergoing treatment, and willingness to undergo rTMS in the future.

##### Credibility/Expectancy

The Credibility Expectancy Questionnaire (CEQ) and the Stanford Expectations of Treatment Scale (SETS) were administered pre- and post-treatment to assess participants’ confidence in the rationale and beliefs about the treatment’s effectiveness.

#### Secondary Outcome Measures

##### Post-Stroke Apathy

Apathy severity was assessed using multiple validated scales, given uncertainty regarding the optimal measurement of apathy in stroke, and to increase sensitivity for treatment effects for use in future research. Our primary outcome was the Lille Apathy Rating Scale (LARS), a 33-item semi-structured interview that captures qualitative features of apathy through respondents’ examples, while maintaining strong validity across neurological populations and excellent discriminant validity relative to depression.^28–30^ Additionally we administered the Apathy Evaluation Scale (AES) given its widespread use in apathy research, facilitating comparison with existing literature;^26^ the Dimensional Apathy Scale (DAS), a 24 item self-rated instrument specifically designed to be independent of motor disability, of particular importance in stroke, and the only scale grounded in the three apathy subtypes (executive, emotional, and initiation);^31,32^ and the apathy subscale of the Frontal Systems Behavior Scale (FrSBe-A), given its strong internal construct validity confirmed through factor analysis,^33,34^ superior reliability in measuring cognitive and behavioral, as opposed to emotional-affective components of apathy,^35^ and convergence with neural markers of frontal dysfunction.^34,36^ Of note, the LARS, AES, DAS, and FrSBe all have demonstrated cross-validity in stroke.^37–40^ Self-rated scales were used alongside informant-rated versions to account for potential differences between participants’ and informants’ perceptions of apathy, particularly in light of potential for anosognosia in stroke.

##### Other Neuropsychiatric symptoms

Measures included the GRID-Hamilton Depression Rating Scale (GRID-HAMD), the Snaith-Hamilton Pleasure Scale (SHAPS), which measures anhedonia, and the PROMIS short-form scales for depression, anxiety, fatigue, and social participation.

##### Quality of life

Measures included the PROMIS short form social participation scale, the Lawton Instrumental Activities of Daily Living Scale (L-IADL), the Brief Inventory of Psychosocial Function (B-IPF), and the Stroke-Specific Quality of Life Scale (SS-QOL). Co-participants’ care burden was additionally measured with the Zarit Burden Interview (ZBI).

### Statistical Analyses

#### Sample Size Justification

As a phase I feasibility study, formal sample-size calculation was not warranted. An enrollment target of 20 participants was selected to meet the primary aims of assessing the safety, feasibility, acceptability, and tolerability of the treatment paradigm, as well as outcome variability for planning future studies and accounting for expected attrition (20%). Statistical analyses were performed in R (version 4.4.2, R Foundation for Statistical Computing, Vienna, Austria) with the rstatix and afex packages. Demographic and clinical variables were summarized using means ± standard deviations or frequencies and percentages. Repeated-measures ANOVAs were conducted to assess omnibus time effects across measures at three time points: at intake, immediate post-treatment, and one-month post-treatment. Significant omnibus effects were then evaluated further via Bonferroni-corrected post-hoc *t-*tests comparing pre-treatment scores with post-treatment scores and pre-treatment scores with one-month follow-up scores using α = 0.025 to correct for multiple comparisons. For each ANOVA, Mauchly’s test was used to assess the assumption of sphericity. For variables that violated this assumption, Greenhouse-Geisser corrected degrees of freedom were used to estimate *p*-values for *F* statistics. Effect sizes for all ANOVAs were estimated using partial ^2^, with effect sizes interpreted using conventional benchmarks as small (0.01-0.059), medium (0.06-0.139), or large (≥ 0.14). Measures sampled at only two time-points (e.g., the NIH toolbox) and post-hoc comparisons for ANOVA effects were conducted using paired-samples *t*-tests, and effect sizes for these differences were reported using Cohen’s *d*, with effect sizes interpreted using conventional benchmarks as small (0.2-0.49), medium (0.50-0.80), or large (≥ 0.80).

## RESULTS

### Participant Recruitment and Characteristics

Participants were enrolled between October 2023 and May 2025. Participant flow and study completion are summarized in the CONSORT diagram (**Figure 2**): 447 records were assessed for eligibility; 83 underwent formal telephone prescreening. Recruitment and enrollment targets were met, with 21 participants enrolled. Following enrollment, five participants were excluded prior to rTMS treatment allocation due to MRI-incompatible materials (n=3); AES < 39 at intake (n=1); new diagnosis of PD (n=1); MRI-related claustrophobia (n=1). 15 participants were then allocated to treatment and began open-label iTBS-rTMS. Among those who started treatment, the retention rate was 93.3%, with 1 participant withdrawing due to a preference for other studies (unrelated to rTMS treatment or study procedures). All 14 remaining participants completed the intervention and were included in the final analysis (**Figure 2**, CONSORT diagram).

**Figure 2:**
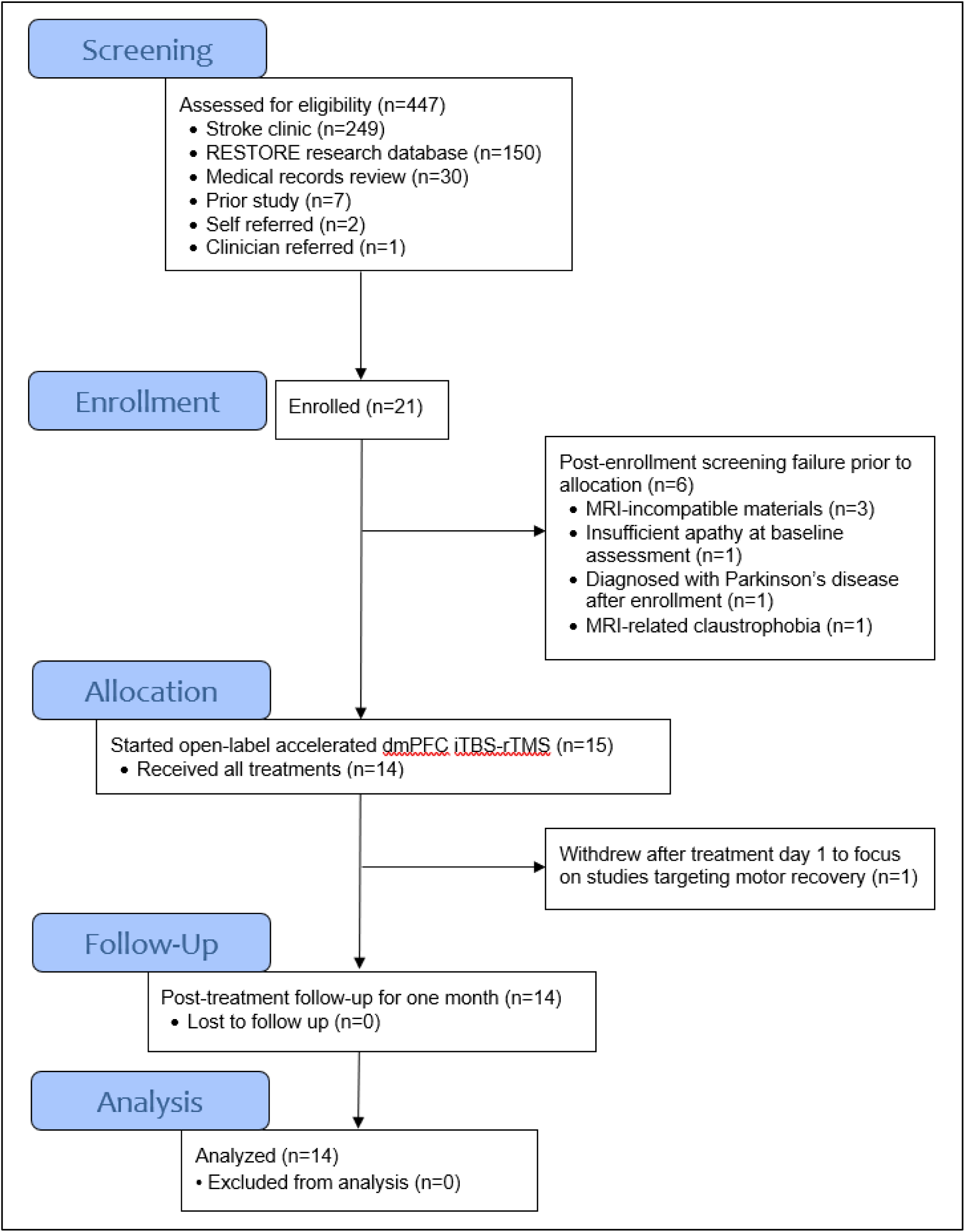
CONSORT Diagram. Flow diagram of participant enrollment and retention.

The baseline demographic characteristics of the 14 participants who underwent rTMS are summarized in **Table 1**. Participants ranged in age from 40 to 80 years (mean ± SD, 61.79 ± 14.03 years), with 11 men and 3 women. The sample was predominantly White (n=11), and the remainder were Black (n=3). Most participants had ischemic strokes (n=12), and the remainder had hemorrhagic strokes (n=2). Lesions were distributed across the left (n=6) and right (n=8) hemispheres (**Table 1, Figure S1**). The mean time since stroke was 55.6 ± 31.6 months (range = 6 to 92 months). Co-participants were primarily spouses (n=8), with others being adult children (n=2), close family members, or friends (n=4).

**Table 1.**
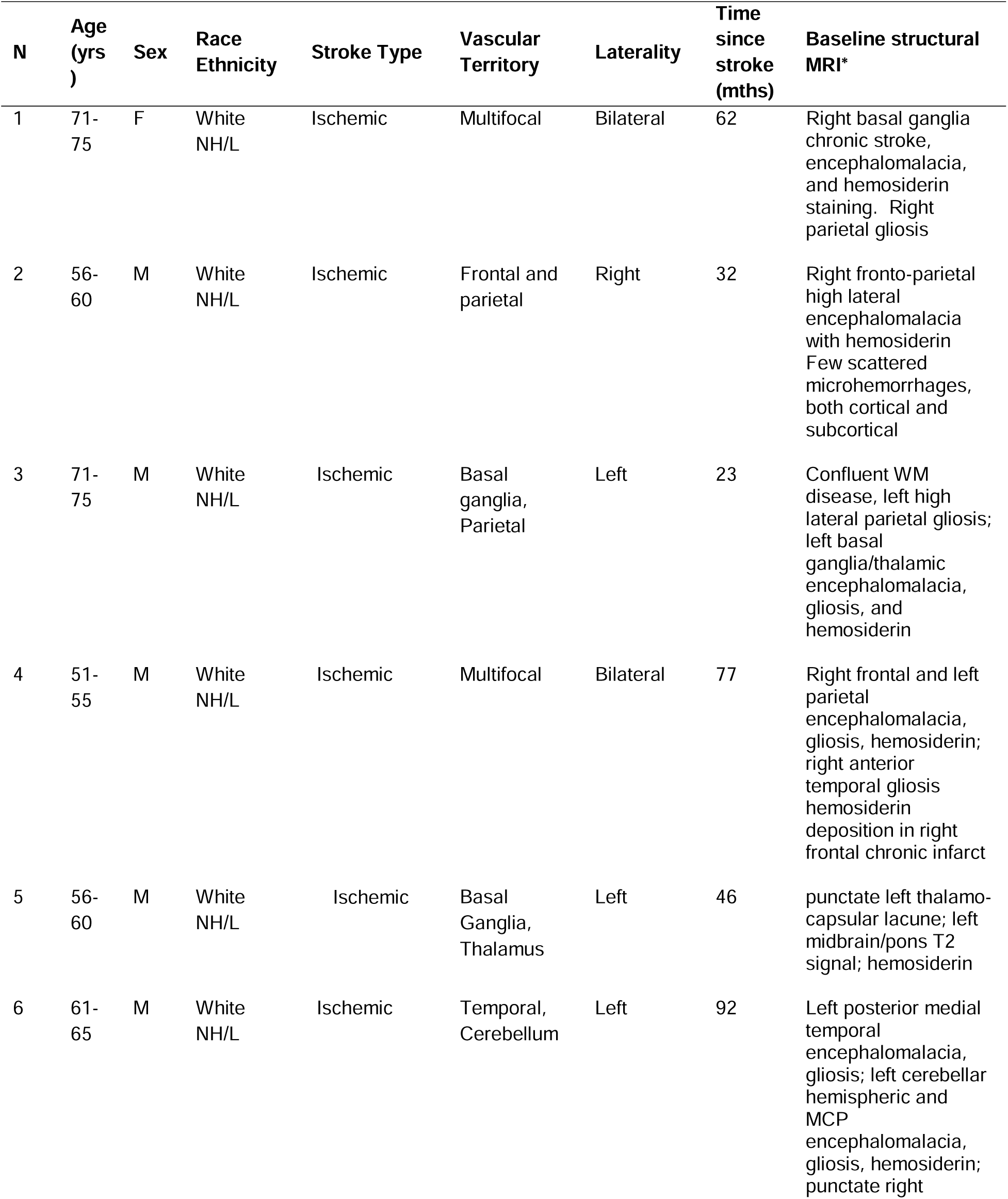
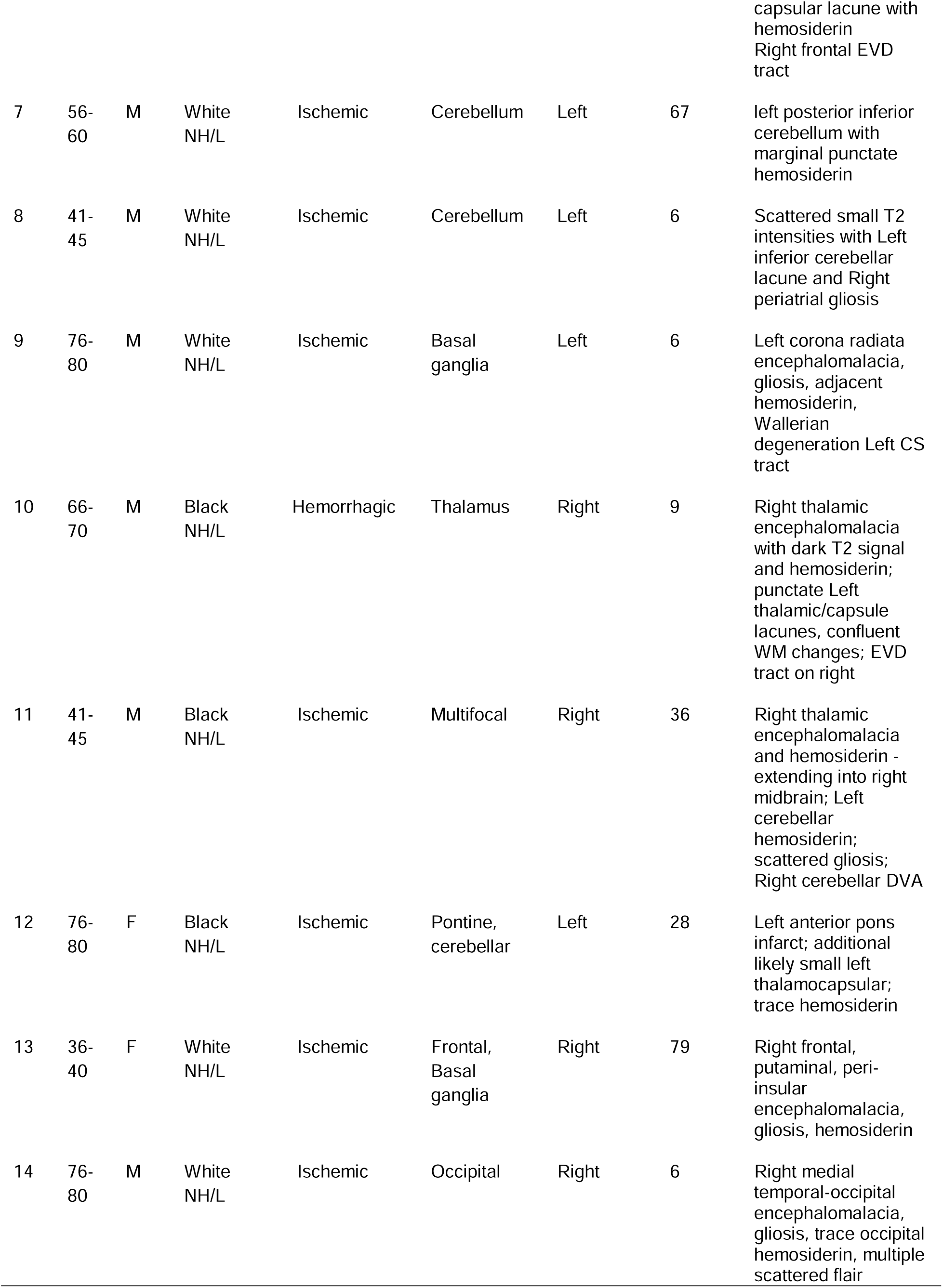
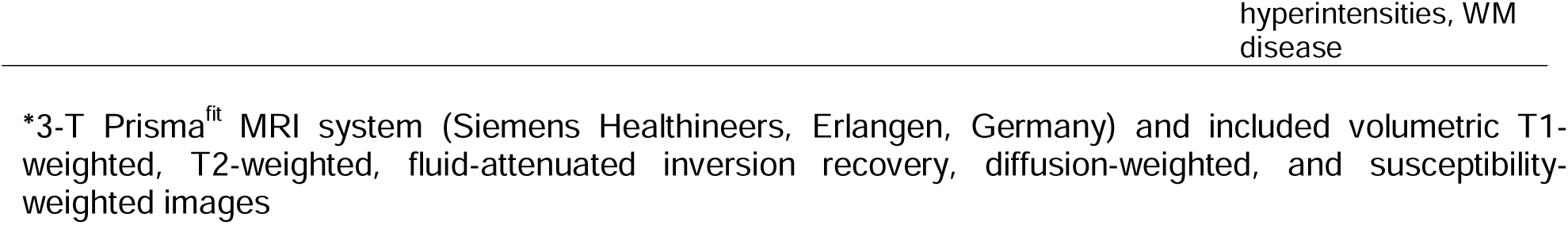
Population characteristics. Baseline demographics and imaging characteristics of all study participants. Abbreviations: N, Participant number; NH/L, Non-Hispanic/Latino; mths, months; MRI, Magnetic Resonance Imaging; M, male; F, female; WM, white matter; MCP, middle cerebellar peduncle; EVD, external ventricular drain; CS, corticospinal; DVA, Developmental venous anomaly

### Primary Outcomes

#### Safety, Feasibility, Tolerability, and Acceptability

No serious adverse events occurred during the study. There were no treatment-related changes in structural MRI scans. One participant demonstrated a 1 mm, clinically silent focus of restricted diffusion without a definite ADC correlate in the right frontal lobe, which was anatomically distant from the site of stimulation and was judged to be unrelated to the intervention (**Figure S2**). Electric field models showed that, on average, participants received 61.3 V/m (47.5-79.9, 10.4) from stimulation within the left dmPFC region of interest. This amount of stimulation is safe and within therapeutic range for effective target engagement (**Figure 3**). There were no decrements or adverse changes in cognitive function or neuropsychiatric symptoms. Participants rated the TMS intervention as highly acceptable and well-tolerated (**Table S1**). Side effects, when reported, were generally mild and improved over the course of treatment with subsequent sessions. (**Figures 4 and S3**). Regarding expectancy effects, the SETS showed that participants had mild positive expectations prior to treatment (mean ± SD, SETS _positive_ 4.31 ± 1.64; SETS _negative_, 2.45 ± 1.34). Regarding treatment credibility, the CEQ reflected that participants viewed the treatment as generally credible pre-treatment (6.05 ± 2.36), and this perception was slightly attenuated post-treatment (5.27 ± 2.27).

**Figure 3:**
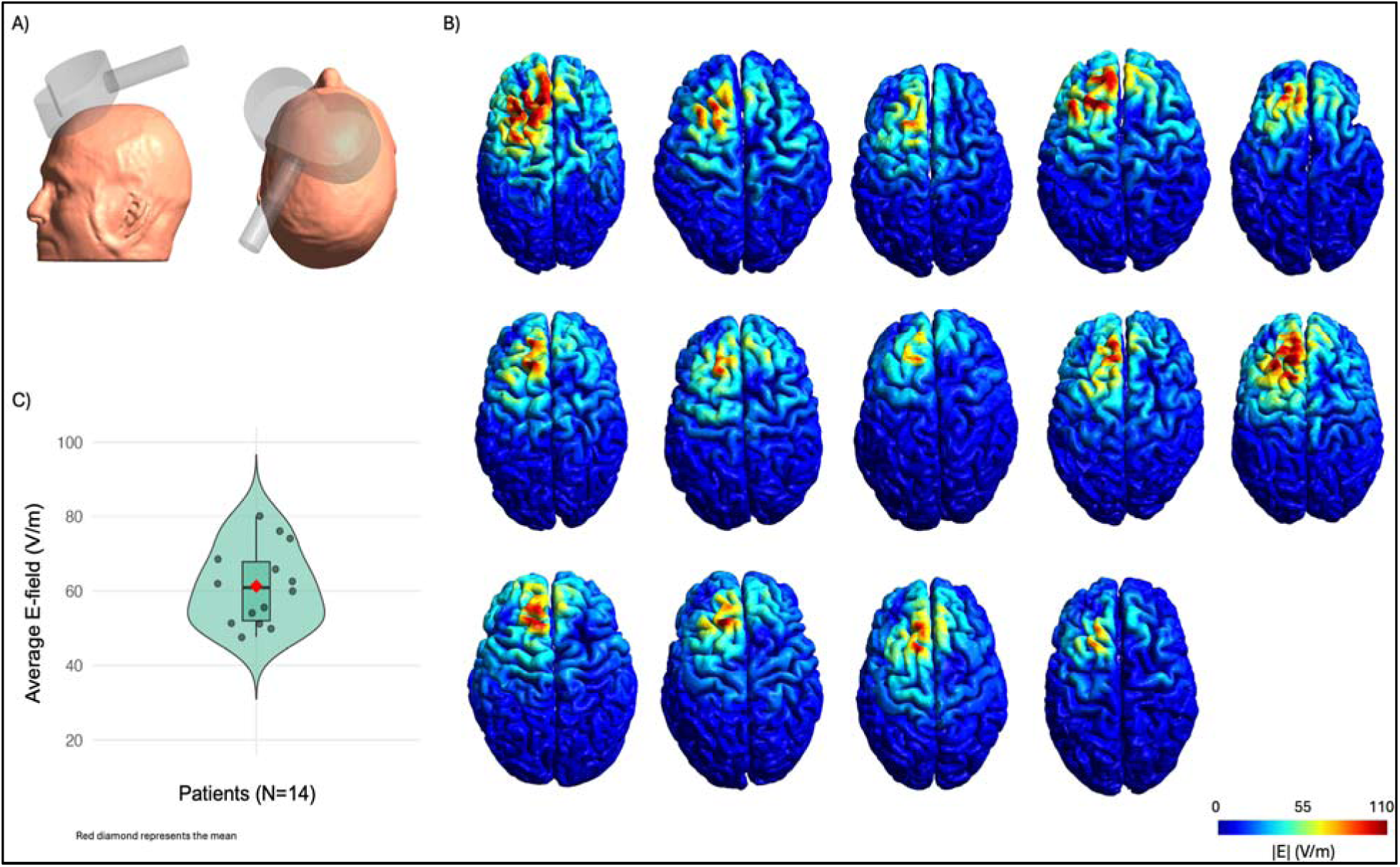
Electric Field Models. To further investigate the amount of 120% rTMS stimulation reaching the target left dmPFC, we performed electric field modeling for each participant. This figure presents A) the position of the stimulation coil over the MNI target (x: -12.63, y: 41.21, and z:75.25) on the standard MNI template head, B) a visualization of electric fields for each of the 14 participants over the head, and C) a boxplot showing the distribution of electric field values per participant. On average, the participants received 61.3 V/m (47.5-79.9, 10.4) from stimulation in the left dmPFC ROI. This amount of stimulation is safe and within therapeutic range for effective target engagement.

**Figure 4:**
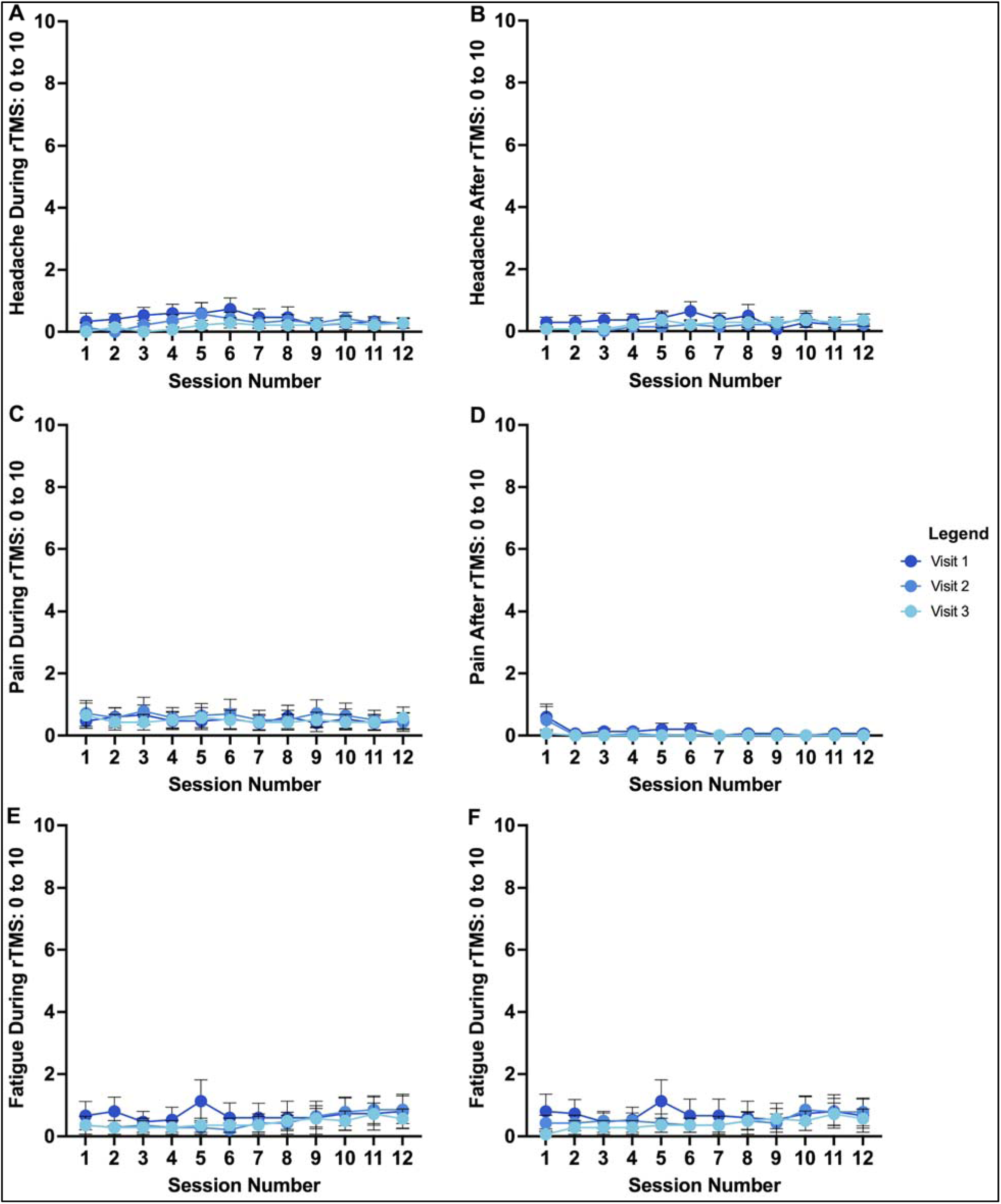
Tolerability – Momentary Assessment. A Momentary Assessment of rTMS was administered after every session on each treatment day (12 sessions per day for 3 days), in which participants rated common rTMS side effects on a 10-point Likert scale (from 0 = not at all to 10 = extremely).

#### Secondary Outcomes

Sample descriptive statistics and inferential statistics for secondary outcomes are presented in **Table 2**. Individual participant and mean changes in these measures over time are also depicted in **Figures 5-6**.

**Figure 5:**
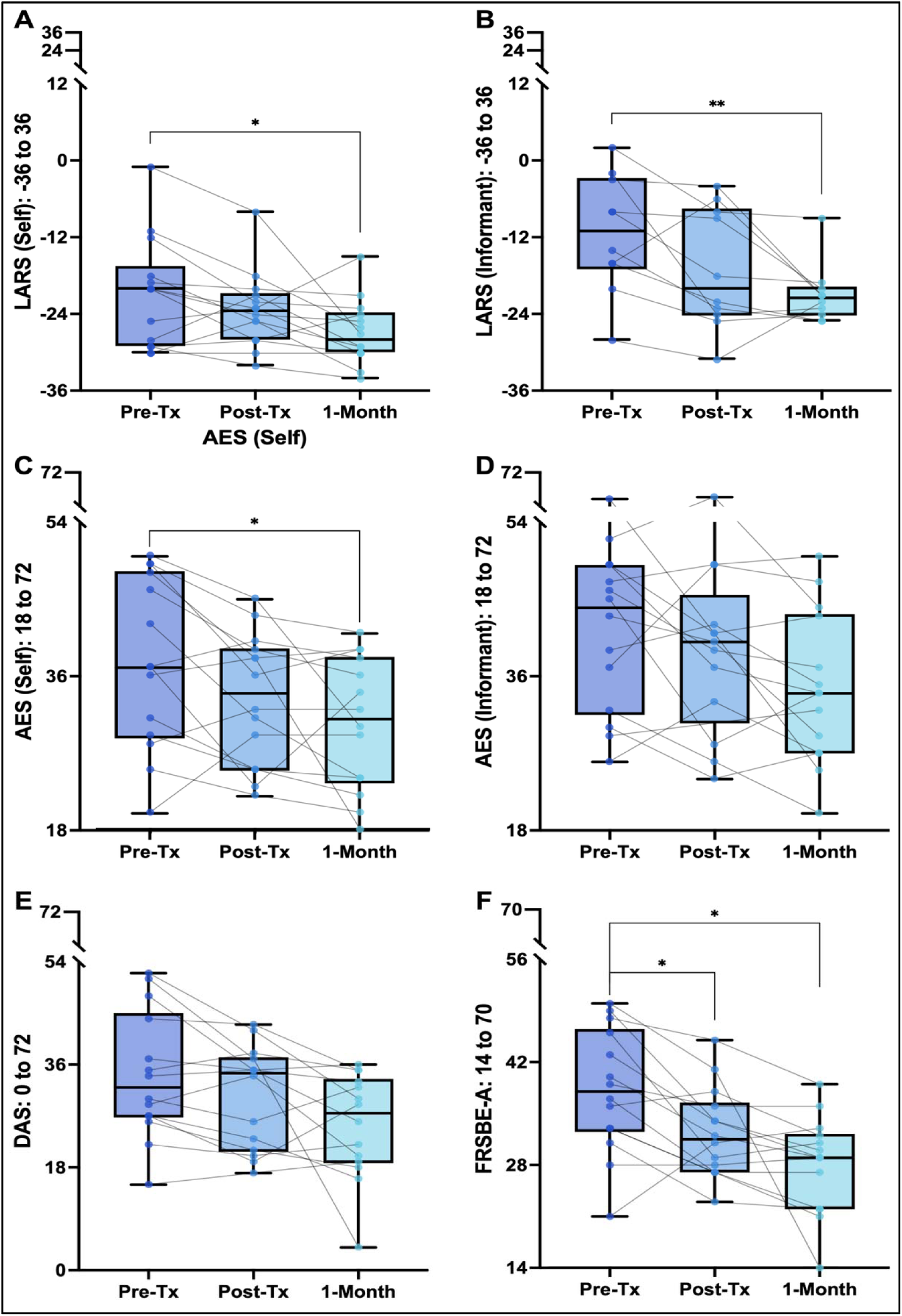
Apathy outcomes across time. Individual trajectories (black lines) and group means ± standard deviations (box plots) for apathy measures at pre-, immediate post-, and one-month treatment. Abbreviations: AES, Apathy Evaluation Scale; LARS, Lille Apathy Rating Scale; DAS, Dimensional Apathy Scale; FrSBe-A, Frontal Systems Behavior Scale–Apathy subscale.

**Figure 6:**
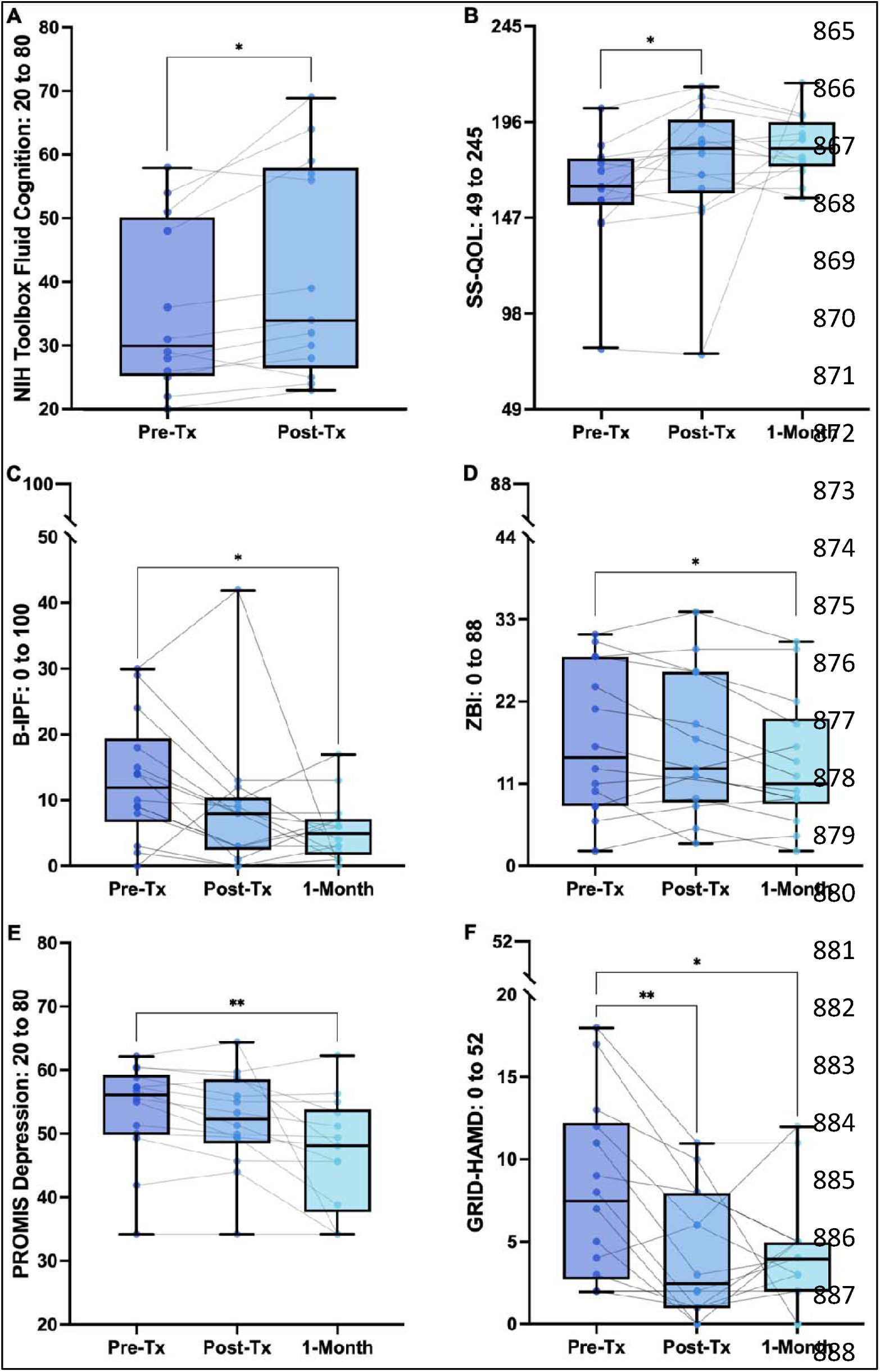
Cognitive, Secondary Neuropsychiatric, and Quality of Life-related outcomes across time. Individual trajectories (black lines) and group means ± standard deviations (box plots) for measures at pre-, immediate post-, and one-month post treatment. Abbreviations: NIHToolbox Fluid Cognition, National Institute of Health Fluid Cognition Composite scores; SS-QOL, Stroke-Specific Quality of Life Scale; B-IPF, Brief Inventory of Psychosocial Functioning; ZBI, Zarit Burden Interview; PROMIS, Patient-Reported Outcomes Measurement Information System short form depression; GRID-HAMD, GRID Hamilton Rating Scale for Depression.

**Table 2:** Secondary Outcomes. Descriptive and inferential statistics, including omnibus time effects and post-hoc comparisons between study time points. Abbreviations: LARS, Lille Apathy Rating Scale; AES, Apathy Evaluation Scale; DAS, Dimensional Apathy Scale; FrSBe-A, Frontal Systems Behavior Scale–Apathy subscale; GRID-HAMD, GRID Hamilton Rating Scale for Depression; PROMIS, Patient-Reported Outcomes Measurement Information System; SHAPS, Snaith-Hamilton Pleasure Scale; MoCA, Montreal Cognitive Assessment; NIH, National Institutes of Health; L-iADL, Lawton Instrumental Activities of Daily Living Scale; B-IPF, Brief Inventory of Psychosocial Functioning; SS-QOL, Stroke-Specific Quality of Life Scale; ZBI, Zarit Burden Interview

|  | Pre-Treatment | Immediate post-treatment | One-Month Follow-Up | Omnibus Difference | Pre-Imm Post Difference | Pre-One Month Follow-Up Difference |
| --- | --- | --- | --- | --- | --- | --- |
| | Mean (SD) | Mean (SD) | Mean (SD) | $F(\eta^2_p)$ | $t$ (Cohen's $d$) | $t$ (Cohen's $d$) |
| <u>*Apathy Measures*</u> |  |  |  |  |  |  |
| LARS | -20.86<br>(8.56) | -23.36<br>(5.89) | -26.86<br>(5.04) | 6.03 (0.32) * | 1.84 (0.49) | 2.92 (0.78) * |
| LARS Informant<sup>†</sup> | -11.30<br>(9.19) | -17.00<br>(9.46) | -20.90<br>(4.65) | 7.40 (0.45) * | 2.19 (0.69) | 4.04 (1.28) ** |
| AES | 37.64<br>(9.97) | 33.29<br>(7.63) | 30.43<br>(7.75) | 5.28 (0.29) * | 2.40 (0.64) ^ | 2.60 (0.69) * |
| AES Informant<sup>‡</sup> | 41.57<br>(9.63) | 39.08<br>(9.89) | 34.57<br>(8.84) | 4.94 (0.29) * | 1.36 (0.38) | 2.51 (0.67) ^ |
| DAS | 34.07<br>(11.19) | 30.36<br>(8.99) | 25.36<br>(9.26) | 5.03 (0.28) * | 1.78 (0.64) ^ | 2.47 (0.66) ^ |
| FrSBe-A | 38.14<br>(8.56) | 32.29<br>(3.14) | 28.14<br>(6.56) | 9.68 (0.43) * | 3.18 (0.85) * | 3.43 (0.92) * |
| <u>*Psychiatric Measures*</u> |  |  |  |  |  |  |
| GRID-HAMD | 8.07<br>(5.48) | 4.21<br>(3.85) | 4.36<br>(3.48) | 7.57 (0.37) * | 4.07 (1.09) ** | 2.83 (0.76) * |
| PROMIS Depression | 53.60<br>(7.78) | 52.11<br>(7.68) | 46.89<br>(8.83) | 7.82 (0.38) * | 2.28 (0.61) ^ | 3.35 (0.90) ** |
| PROMIS Anxiety | 52.40<br>(8.38) | 51.84<br>(8.83) | 49.56<br>(9.74) | 1.29 (0.09) | 0.31 (0.08) | 1.68 (0.45) |
| PROMIS Fatigue | 53.86<br>(7.42) | 52.85<br>(8.12) | 49.33<br>(10.32) | 2.26 (0.15) | 0.77 (0.21) | 1.66 (0.45) |
| PROMIS Social Participation | 48.39<br>(6.72) | 49.24<br>(7.51) | 52.69<br>(5.45) | 2.15 (0.14) | 0.61 (0.16) | 1.89 (0.50) |
| SHAPS | 2.14<br>(2.93) | 1.79<br>(3.33) | 0.71<br>(1.27) | 1.68 (0.11) | 0.57 (0.15) | 1.68 (0.45) |
| <u>*Cognitive Function*</u> |  |  |  |  |  |  |
| MoCA | 24.50<br>(3.23) | 25.00<br>(3.67) | 25.20<br>(3.56) | 0.65 (0.05) | 0.76 (0.21) | 1.01 (0.28) |
| NIH Toolbox Cognition Fluid<sup>¥</sup> | 35.67<br>(13.44) | 40.25<br>(16.92) | - | - | 2.67 (0.77) * | - |
| NIH Toolbox Cognition Crystal<sup>¥</sup> | 54.92<br>(9.16) | 55.33<br>(7.88) | - | - | 0.26 (0.08) | - |
| NIH Toolbox Cognition Total<sup>¥</sup> | 44.25<br>(12.12) | 47.42<br>(12.10) | - | - | 2.15 (0.62) ^ | - |
| PROMIS Cognition | 44.84<br>(6.59) | 46.46<br>(5.43) | 48.92<br>(6.93) | 1.23 (0.09) | 1.47 (0.39) | 1.27 (0.34) |
| <u>*Quality of Life*</u> |  |  |  |  |  |  |
| L-iADL | 5.50<br>(2.47) | 5.36<br>(2.34) | 5.43<br>(2.38) | 0.13 (0.01) | 0.56 (0.15) | 0.23 (0.06) |
| B-IPF | 13.21<br>(9.42) | 8.57<br>(10.56) | 5.57 (4.78) | 4.27 (0.25) * | 2.08 (0.56) | 3.32 (0.89) * |
| SS-QOL | 161.21<br>(28.15) | 174.14<br>(34.28) | 183.43<br>(16.04) | 3.24 (0.20) ^ | 2.66 (0.71) * | 2.33 (0.62) ^ |
| ZBI ‡ | 16.86<br>(9.93) | 16.38<br>(9.77) | 13.64<br>(8.66) | 5.11 (0.30) * | 0.75 (0.21) | 2.70 (0.72) * |

ANOVA significance level calculated with Greenhouse Geisser degrees of freedom when sphericity assumption was violated.
N=14 for all analyses unless otherwise specified; ‡-n=13 at post; †-n=10; ¥-n=12
\*- $p < 0.05$ ( $p < 0.025$ for post-hoc comparisons); ^- $p < 0.10$ ( $0.025 < p < 0.05$ for post-hoc comparisons)

#### Apathy

All apathy measures (self- and informant-rated) showed statistically significant omnibus changes over time, with large effect sizes (η _p_^2^ > 0.14; *p* < 0.05 for all measures: AES, LARS, DAS, FrSBe-A). Post-hoc tests revealed that apathy was significantly reduced for all included measures pretreatment-to-one month follow up (AES: 37.64 ± 9.97 to 30.43 ± 7.75; LARS: - 20.86 ± 8.56 to -26.86 ± 5.04; LARS informant: -11.30 ± 9.19 to -20.90 ± 4.65; FrsBe-A: 38.14 ± 8.56 to 28.14 ± 6.56; *p* < 0.025 for all) excluding the AES informant (41.57 ± 9.63 to 34.57 ± 8.84, *p* = 0.026) and DAS scales (34.07 ± 11.19 to 25.36 ± 9.26*, p* =0.028) with medium-to-large effect sizes (*d* = 0.66-1.28). However, post-hoc tests showed less consistent effects on apathy reduction from pre- to immediate post-treatment, suggesting that some therapeutic effects accumulated over the weeks following stimulation. Specifically, there were significant reductions from pre- to immediate post-treatment for the AES (37.64 ± 9.97 to 33.29 ± 7.63; *p* = 0.032), DAS (34.07 ± 11.19 to 30.36 ± 8.99; *p* = 0.033) and FrSBe-A (38.14 ± 8.56 to 32.29 ± 3.14; *p* = 0.015) with medium-to-large effect sizes (*d =* 0.64-0.85, *p* < 0.05 for all). In contrast, the AES informant, LARS, and the LARS informant scales exhibited non-significant reductions with small-to-medium effect sizes (*d* = 0.38-0.69) from pre- to immediate post-treatment. Subscale analysis for the LARS, AES, DAS, and FrSBe is reported in **Table S2.**

#### Cognitive Function

The fluid cognition scale from the NIH toolbox exhibited a significant increase from pre- to post-treatment (35.67 ± 13.44 to 40.25 ± 16.92, *p* < 0.05).

#### Quality of Life

The B-IPF and the ZBI both exhibited significant omnibus change with large effect sizes of improvement (η _p_ ^2^ > 0.14; *p* < 0.05). The SS-QOL exhibited a marginally significant (*p* < 0.10) omnibus change with a large effect size (η _p_^2^ > 0.14). Post-hoc tests revealed that the B-IPF and the ZBI both improved significantly (*p* < 0.025) at the one-month follow-up relative to pre-treatment, with medium-to-large effects (*d* = 0.72 and 0.89, respectively), but showed no significant change between pre- and post-treatment. Conversely, the SS-QOL exhibited significant (*p* < 0.025) improvement from pre- to post-treatment, with a medium effect size (*d* = 0.71), and a marginally significant improvement (*p* < 0.05) at the one-month follow-up time point with a medium effect size (*d* = 0.62).

#### Other Neuropsychiatric symptoms

Depression symptoms were mild-to-absent at baseline (pre-treatment GRID-HAMD, 8.07 ± 5.48). Nevertheless, there were statistically significant omnibus changes with large effect sizes on the reduction of depression, as measured by the GRID-HAMD (η_p_^2^ = 0.37) and PROMIS short-form depression scale (η_p_^2^ = 0.38). Post-hoc tests revealed that depression scores were significantly lower at post-treatment for the HAMD (and marginally lower for PROMIS-Depression [*p* = 0.04]), and both measures were significantly lower at the one-month follow-up with medium-to-large effect sizes (HAMD *d* = 0.76, PROMIS-Depression *d* = 0.90). No other psychiatric measures exhibited statistically significant omnibus changes; omnibus effect-size estimates were large (η_p_^2^ > 0.14) for the PROMIS short-form fatigue and social participation scales.

## Discussion

The results of this open-label phase I study suggest that accelerated iTBS-rTMS targeting the left dmPFC is safe, feasible, well-tolerated, and highly acceptable in stroke survivors with clinically significant apathy. With 21,600 total iTBS pulses delivered across 36 sessions over 3 treatment days, this protocol represents one of the highest dose accelerated rTMS regimens evaluated to our knowledge in a chronic stroke population to date. No serious adverse events occurred, and side effects, when they occurred, were generally rated as tolerable and improving over the course of treatment. No treatment-related changes were observed on neuroradiological scanning. The inclusion of multimodal MRI sequences obtained both before and after treatment further strengthens the safety assessment of this study and builds upon prior rTMS literature, which has often relied on more limited anatomical imaging. Cognitive performance remained stable or improved modestly during the follow-up period. The study met all recruitment, enrollment, and retention targets, with 93.3% retention among participants allocated to treatment and no withdrawals attributable to treatment or study procedures.

Despite not being powered for efficacy, we observed significant reductions in apathy across several self- and informant-rated measures, with medium-to-large effect sizes across all scales. Improvements were most pronounced at one month following treatment, in line with the broader TMS literature suggesting clinical benefit often continues to accumulate in the weeks following stimulation.^41,42^

By 1-month follow-up, mean scores on all apathy measures with published clinical thresholds (LARS, AES, DAS) fell below the cutoffs for clinically significant apathy.^25,32,43^ For the AES, observed mean improvements of 7.2 (self-rated) and 7.0 (informant-rated) exceeded the anchor-based minimum clinically important difference (MCID) of approximately 3.3.^44^ For the LARS, mean reductions of 6.0 points (self-rated) and 9.6 points (informant-rated) corresponded to a shift of at least one apathy severity category.^28^ While thresholds for MCID or severity categories are not well-established for the DAS or FrSBe-A, these measures demonstrated comparable magnitudes of improvement, with moderate-to-large effect sizes.

Reductions in apathy were accompanied by parallel improvement in psychosocial functioning, co-participant care burden, and clinically meaningful improvement in stroke-specific quality of life.^45^ These convergent findings across multiple domains reduce the likelihood that results reflect measurement artifact alone and instead suggest meaningful clinical behavioral change. Though we saw statistically significant improvement in depression, scores on the GRID-HAMD and SHAPS were in the normal to mild ranges at baseline,^46,47^ indicating pre-treatment minimal to absent depressive symptoms in the sample. Additionally, given that anhedonia and PSA share common behavioral symptoms (e.g., reduced energy and loss of motivation), reductions in depression scores likely reflect decreases in overlapping apathy and anhedonia symptoms. Consistent with this idea, the SHAPS – a scale designed primarily to capture the hedonic pleasure aspect of anhedonia rather than loss of energy or motivation – showed no significant change over the course of the study.

### Targeting motivational circuitry after stroke

This study extends prior neuromodulation work in stroke by shifting the therapeutic focus from motor and language recovery to the neuropsychiatric sequelae that often limit rehabilitation engagement and long-term functional outcomes.^48^ Whereas most rTMS protocols have targeted the dlPFC based on depression treatment models, PSA is a distinct neuropsychiatric syndrome with meaningfully distinguishable neuroanatomic correlates. The dmPFC, through its dense interconnections with the ACC and ventral striatum, occupies a central position in the motivational network that integrates cognitive, affective, and motor inputs necessary to drive goal-directed behavior. By engaging this circuit, the present intervention may engage downstream networks critical for motivated behaviors. Importantly, our findings complement and extend a prior trial of dmPFC rTMS for PSA, demonstrating that high-dose accelerated delivery is not only feasible but may amplify clinical response.^22^

### Clinical Implications

Apathy is a major impediment to recovery after stroke because it directly undermines participation in rehabilitation and adaptive compensatory behaviors. An intervention capable of reducing apathy in a brief treatment window could significantly enhance rehabilitation engagement and bolster existing interventions targeting functional recovery. In this study, the intervention appeared highly manageable to participants, as reflected by a 93.3% completion rate among participants who initiated rTMS treatment. By pioneering an accelerated rTMS paradigm with 12 sessions of iTBS per day and relatively short intersession intervals of 10-15 minutes, the present treatment also compressed each treatment day to approximately 3 hours, which compares favorably to common accelerated rTMS approaches that can last up to 10 hours.^49^ We believe that such a compressed treatment paradigm is well-suited to the chronic stroke population and could facilitate integration with standard rehabilitation interventions such as cognitive, speech, physical, or occupational therapies. In contrast, other accelerated neuromodulation paradigms requiring longer intersession intervals require substantially longer treatment days, limiting their feasibility in neurologic populations.^49^

### Limitations

Several limitations warrant consideration. First, the single-arm, open-label design precludes causal inference and raises the possibility of expectancy effects; however, SETS and CEQ scores indicated only modest positive expectancy at baseline, and perceived treatment credibility did not increase following intervention. Moreover, peak effects arising at 1-month follow-up would be atypical for placebo effects, and the inclusion of informant-rated assessments in the present study provides a collateral reference that is at least partially independent of self-report. Second, the small sample size and demographic homogeneity limit generalizability and may inflate standard effect size estimates, though effect sizes were consistent across multiple independent measures and informants, supporting the strength of the signal. Third, although heterogeneous stroke lesion profiles enhance ecological validity, they may also preclude definitive conclusions regarding structure-function relationships, though PSA does not appear to depend on stroke type, location, or laterality, lending evidence for dysfunction in a common network underlying apathy and supporting a transdiagnostic treatment approach. Finally, follow-up was limited to one month, with ongoing, accumulating benefits at the final time-point across several measures; thus, the absolute magnitude of delayed benefits and treatment durability remains unknown, though the observed trajectory is consistent with prior brain stimulation studies showing delayed clinical response.^41,42^

### Future directions

These findings provide strong justification for a randomized, sham-controlled trial to confirm the efficacy of accelerated left dmPFC iTBS-rTMS for PSA, and to further identify predictors of response. By reporting all dosing parameters, we hope to facilitate reproducibility and encourage further research that builds on these results. We suggest that studies should incorporate biomarkers of effective target engagement, such as functional connectivity, TMS-EEG, and effort-based behavioral tasks, and examine whether combining brain stimulation with other behavioral activation and rehabilitation therapies produces synergistic gains. Given the multidimensional nature of apathy, future studies may also explore additional neuroanatomical targets to elucidate differential effects across its subdomains.^14^ Longer follow-up intervals and booster paradigms may also help to further establish optimal treatment parameters and maintenance strategies. Finally, individualized connectivity-based targeting may further optimize therapeutic precision, especially in stroke, where post-infarct reorganization can alter functional network architecture and limit the accuracy of conventional targeting approaches.

## Conclusions

Accelerated left dmPFC-targeted iTBS-rTMS appears to be safe, feasible, and highly acceptable in chronic stroke survivors with clinically significant apathy. Treatment was associated with significant improvements in self- and informant-rated apathy, as well as related psychosocial outcomes. By directly engaging the neuroanatomical substrates of apathy and minimizing treatment burden, this paradigm has the potential not only to reduce apathy but also to improve long-term functional outcomes in stroke. These preliminary data support further investigation into circuit-based neuromodulation as a strategy to restore motivation and enhance rehabilitation engagement, thereby improving long-term recovery after stroke. This paradigm may further lay groundwork for a next generation of versatile, non-invasive, mechanism-driven treatment paradigms with application across transdiagnostic neuropsychiatric syndromes and clinically diverse populations.

## Acknowledgements

This project was supported, in part, by the National Institute of General Medical Sciences of the National Institutes of Health under grant number P30GM154630, as well as the National Institute of Child Health and Human Development under grant number P50HD118628. The content is solely the responsibility of the authors and does not necessarily represent the official views of the National Institutes of Health.

The authors also acknowledge Drs. Federico Rodriguez-Porcel, MD, and Ezequiel Gleichgerrcht, MD, PhD, for their assistance.

## Sources of Funding

Research reported in this publication was supported by pilot funding from the National Institutes of Health National Center of Neuromodulation for Rehabilitation (NC NM4R) under Grant Number 5P2CHD086844 and the National Institute on Drug Abuse under Grant Number R25DA020537.

## Disclosure

None for all authors

## Notes

### Competing Interest Statement

The authors have declared no competing interest.

### Clinical Trial

NCT0587845

### Author Declarations

The study was conducted at the Medical University of South Carolina (MUSC), a comprehensive stroke center in South Carolina and was approved by the MUSC Institutional Review Board (IRB; Pro00126436).

## References

1. Hackett ML, Kohler S, O’Brien JT, Mead GE. Neuropsychiatric outcomes of stroke. Lancet Neurol. 2014;13:525–534. doi: 10.1016/S1474-4422(14)70016-X

2. Robert P, Lanctot KL, Aguera-Ortiz L, Aalten P, Bremond F, Defrancesco M, Hanon C, David R, Dubois B, Dujardin K, et al. Is it time to revise the diagnostic criteria for apathy in brain disorders? The 2018 international consensus group. Eur Psychiatry. 2018;54:71–76. doi: 10.1016/j.eurpsy.2018.07.008

3. Brodaty H, Liu Z, Withall A, Sachdev PS. The longitudinal course of post-stroke apathy over five years. J Neuropsychiatry Clin Neurosci. 2013;25:283–291. doi: 10.1176/appi.neuropsych.12040080

4. Douven E, Kohler S, Schievink SHJ, van Oostenbrugge RJ, Staals J, Verhey FRJ, Aalten P. Baseline Vascular Cognitive Impairment Predicts the Course of Apathetic Symptoms After Stroke: The CASPER Study. Am J Geriatr Psychiatry. 2018;26:291–300. doi: 10.1016/j.jagp.2017.09.022

5. Hollocks MJ, Lawrence AJ, Brookes RL, Barrick TR, Morris RG, Husain M, Markus HS. Differential relationships between apathy and depression with white matter microstructural changes and functional outcomes. Brain. 2015;138:3803–3815. doi: 10.1093/brain/awv304

6. Mayo NE, Fellows LK, Scott SC, Cameron J, Wood-Dauphinee S. A longitudinal view of apathy and its impact after stroke. Stroke. 2009;40:3299–3307. doi: 10.1161/STROKEAHA.109.554410

7. Santa N, Sugimori H, Kusuda K, Yamashita Y, Ibayashi S, Iida M. Apathy and functional recovery following first-ever stroke. Int J Rehabil Res. 2008;31:321–326. doi: 10.1097/MRR.0b013e3282fc0f0e

8. Eurelings LS, van Dalen JW, Ter Riet G, Moll van Charante EP, Richard E, van Gool WA, Almeida OP, Alexandre TS, Baune BT, Bickel H, et al. Apathy and depressive symptoms in older people and incident myocardial infarction, stroke, and mortality: a systematic review and meta-analysis of individual participant data. Clin Epidemiol. 2018;10:363–379. doi: 10.2147/CLEP.S150915

9. Tay J, Morris RG, Tuladhar AM, Husain M, de Leeuw FE, Markus HS. Apathy, but not depression, predicts all-cause dementia in cerebral small vessel disease. J Neurol Neurosurg Psychiatry. 2020;91:953–959. doi: 10.1136/jnnp-2020-323092

10. Ruiz-Franco ML, Amaya-Pascasio L, Gil-Rodriguez M, Arjona-Padillo A, Garcia-Pinteno J, Rodriguez-Sanchez AJ, Sanchez-Kuhn A, Flores P, Martinez-Sanchez P. Treatment of apathy in stroke patients: a systematic review. Front Neurol. 2025;16:1702325. doi: 10.3389/fneur.2025.1702325

11. Hijikuro S, Kaneko T, Ikeda K. Interventions for post-stroke apathy and their effects: A scoping review. Clin Rehabil. 2025;39:1425–1437. doi: 10.1177/02692155251374911

12. McNamara ML, Dalton SGH. A scoping review of available interventions to improve transportation access for chronic stroke survivors. Journal of Transport & Health. 2024;37:101820. doi: 10.1016/j.jth.2024.101820

13. Le Heron C, Apps MAJ, Husain M. The anatomy of apathy: A neurocognitive framework for amotivated behaviour. Neuropsychologia. 2018;118:54–67. doi: 10.1016/j.neuropsychologia.2017.07.003

14. Levy R, Dubois B. Apathy and the functional anatomy of the prefrontal cortex-basal ganglia circuits. Cereb Cortex. 2006;16:916–928. doi: 10.1093/cercor/bhj043

15. Steffens DC, Fahed M, Manning KJ, Wang L. The neurobiology of apathy in depression and neurocognitive impairment in older adults: a review of epidemiological, clinical, neuropsychological and biological research. Transl Psychiatry. 2022;12:525. doi: 10.1038/s41398-022-02292-3

16. Lavin C, Melis C, Mikulan E, Gelormini C, Huepe D, Ibanez A. The anterior cingulate cortex: an integrative hub for human socially-driven interactions. Front Neurosci. 2013;7:64. doi: 10.3389/fnins.2013.00064

17. Tay J, Lisiecka-Ford DM, Hollocks MJ, Tuladhar AM, Barrick TR, Forster A, O’Sullivan MJ, Husain M, de Leeuw FE, Morris RG, et al. Network neuroscience of apathy in cerebrovascular disease. Prog Neurobiol. 2020;188:101785. doi: 10.1016/j.pneurobio.2020.101785

18. George MS, Lisanby SH, Sackeim HA. Transcranial magnetic stimulation: applications in neuropsychiatry. Arch Gen Psychiatry. 1999;56:300–311. doi: 10.1001/archpsyc.56.4.300

19. Beynel L, Powers JP, Appelbaum LG. Effects of repetitive transcranial magnetic stimulation on resting-state connectivity: A systematic review. Neuroimage. 2020;211:116596. doi: 10.1016/j.neuroimage.2020.116596

20. Caulfield KA, Fleischmann HH, George MS, McTeague LM. A transdiagnostic review of safety, efficacy, and parameter space in accelerated transcranial magnetic stimulation. J Psychiatr Res. 2022;152:384–396. doi: 10.1016/j.jpsychires.2022.06.038

21. Somaa FA, de Graaf TA, Sack AT. Transcranial Magnetic Stimulation in the Treatment of Neurological Diseases. Front Neurol. 2022;13:793253. doi: 10.3389/fneur.2022.793253

22. Sasaki N, Hara T, Yamada N, Niimi M, Kakuda W, Abo M. The Efficacy of High-Frequency Repetitive Transcranial Magnetic Stimulation for Improving Apathy in Chronic Stroke Patients. Eur Neurol. 2017;78:28–32. doi: 10.1159/000477440

23. Padala PR, Padala KP, Lensing SY, Jackson AN, Hunter CR, Parkes CM, Dennis RA, Bopp MM, Caceda R, Mennemeier MS, et al. Repetitive transcranial magnetic stimulation for apathy in mild cognitive impairment: A double-blind, randomized, sham-controlled, cross-over pilot study. Psychiatry Res. 2018;261:312–318. doi: 10.1016/j.psychres.2017.12.063

24. Padala PR, Boozer EM, Lensing SY, Parkes CM, Hunter CR, Dennis RA, Caceda R, Padala KP. Neuromodulation for Apathy in Alzheimer’s Disease: A Double-Blind, Randomized, Sham-Controlled Pilot Study. J Alzheimers Dis. 2020;77:1483–1493. doi: 10.3233/JAD-200640

25. Lueken U, Evens R, Balzer-Geldsetzer M, Baudrexel S, Dodel R, Graber-Sultan S, Hilker-Roggendorf R, Kalbe E, Kaut O, Mollenhauer B, et al. Psychometric properties of the apathy evaluation scale in patients with Parkinson’s disease. Int J Methods Psychiatr Res. 2017;26. doi: 10.1002/mpr.1564

26. Marin RS, Biedrzycki RC, Firinciogullari S. Reliability and validity of the Apathy Evaluation Scale. Psychiatry Res. 1991;38:143–162. doi: 10.1016/0165-1781(91)90040-v

27. Scrivener CL, Reader AT. Variability of EEG electrode positions and their underlying brain regions: visualizing gel artifacts from a simultaneous EEG-fMRI dataset. Brain Behav. 2022;12:e2476. doi: 10.1002/brb3.2476

28. Sockeel P, Dujardin K, Devos D, Deneve C, Destee A, Defebvre L. The Lille apathy rating scale (LARS), a new instrument for detecting and quantifying apathy: validation in Parkinson’s disease. J Neurol Neurosurg Psychiatry. 2006;77:579–584. doi: 10.1136/jnnp.2005.075929

29. Dujardin K, Sockeel P, Delliaux M, Destee A, Defebvre L. The Lille Apathy Rating Scale: validation of a caregiver-based version. Mov Disord. 2008;23:845–849. doi: 10.1002/mds.21968

30. Fernandez-Matarrubia M, Matias-Guiu JA, Moreno-Ramos T, Valles-Salgado M, Marcos-Dolado A, Garcia-Ramos R, Matias-Guiu J. Validation of the Lille’s Apathy Rating Scale in Very Mild to Moderate Dementia. Am J Geriatr Psychiatry. 2016;24:517–527. doi: 10.1016/j.jagp.2015.09.004

31. Radakovic R, Abrahams S. Developing a new apathy measurement scale: Dimensional Apathy Scale. Psychiatry Res. 2014;219:658–663. doi: 10.1016/j.psychres.2014.06.010

32. Santangelo G, D’Iorio A, Piscopo F, Cuoco S, Longo K, Amboni M, Baiano C, Tafuri D, Pellecchia MT, Barone P, et al. Assessment of apathy minimising the effect of motor dysfunctions in Parkinson’s disease: a validation study of the dimensional apathy scale. Qual Life Res. 2017;26:2533–2540. doi: 10.1007/s11136-017-1569-6

33. Stout JC, Ready RE, Grace J, Malloy PF, Paulsen JS. Factor analysis of the frontal systems behavior scale (FrSBe). Assessment. 2003;10:79–85. doi: 10.1177/1073191102250339

34. Carvalho JO, Ready RE, Malloy P, Grace J. Confirmatory factor analysis of the Frontal Systems Behavior Scale (FrSBe). Assessment. 2013;20:632–641. doi: 10.1177/1073191113492845

35. Lane-Brown AT, Tate RL. Measuring apathy after traumatic brain injury: Psychometric properties of the Apathy Evaluation Scale and the Frontal Systems Behavior Scale. Brain Inj. 2009;23:999–1007. doi: 10.3109/02699050903379347

36. Wong KS, Chou T, Peters AT, Ellard KK, Nierenberg AA, Dougherty DD, Deckersbach T. Convergence between behavioral, neural, and self-report measures of cognitive control: The Frontal Systems Behavior Scale in bipolar disorder. J Psychiatr Res. 2022;150:317–323. doi: 10.1016/j.jpsychires.2022.03.053

37. Jamali A, Baluchnejadmojarad T, Jazaeri SZ, Abedi S, Mehdizadeh H, Taghizadeh G. Lille Apathy Rating Scale-Patient Version in Stroke Survivors: Psychometric Properties and Diagnostic Accuracy. J Am Med Dir Assoc. 2024;25:105193. doi: 10.1016/j.jamda.2024.105193

38. Jamali A, Baluchnejadmojarad T, Jazaeri SZ, Abedi S, Mehdizadeh H, Sharabiani PTA, Taghizadeh G. Reliability, validity, and diagnostic accuracy of the apathy evaluation scale in chronic stroke survivors. BMC Psychiatry. 2025;25:201. doi: 10.1186/s12888-025-06626-5

39. Jamali A, Baluchnejadmojarad T, Radakovic R, Jazaeri SZ, Mehdizadeh H, Taghizadeh G. Validation of the dimensional apathy scale and predictors of apathy in stroke survivors. Sci Rep. 2025;15:39949. doi: 10.1038/s41598-025-23757-7

40. Caracuel A, Verdejo-Garcia A, Fernandez-Serrano MJ, Moreno-Lopez L, Santago-Ramajo S, Salinas-Sanchez I, Perez-Garcia M. Preliminary validation of the Spanish version of the Frontal Systems Behavior Scale (FrSBe) using Rasch analysis. Brain Inj. 2012;26:844–852. doi: 10.3109/02699052.2012.655365

41. Apostol M, Valles T, Corlier J, Leuchter M, Young A, Artin H, Koek R, Einstein E, Wilke S, Tadayonnejad R, et al. Efficacy of 5×5 accelerated versus conventional repetitive transcranial magnetic stimulation (rTMS) for treatment-resistant depression. Res Sq. 2025. doi: 10.21203/rs.3.rs-7377114/v1

42. Koerselman F, Laman DM, van Duijn H, van Duijn MA, Willems MA. A 3-month, follow-up, randomized, placebo-controlled study of repetitive transcranial magnetic stimulation in depression. J Clin Psychiatry. 2004;65:1323–1328. doi: 10.4088/jcp.v65n1005

43. Zahodne LB, Young S, Kirsch-Darrow L, Nisenzon A, Fernandez HH, Okun MS, Bowers D. Examination of the Lille Apathy Rating Scale in Parkinson disease. Mov Disord. 2009;24:677–683. doi: 10.1002/mds.22441

44. Tumati S, Herrmann N, Perin J, Rosenberg PB, Lerner AJ, Mintzer J, Padala PR, Brawman-Mintzer O, van Dyck CH, Porsteinsson AP, et al. Measuring clinically relevant change in apathy symptoms in ADMET and ADMET 2. Int Psychogeriatr. 2024;36:1232–1244. doi: 10.1017/S1041610224000711

45. Wong GK, Lee A, Wong A, Ho FL, Leung SL, Zee BC, Poon WS, Siu DY, Abrigo JM, Mok VC. Clinically important difference of Stroke-Specific Quality of Life Scale for aneurysmal subarachnoid hemorrhage. J Clin Neurosci. 2016;33:209–212. doi: 10.1016/j.jocn.2016.05.029

46. Williams JB, Kobak KA, Bech P, Engelhardt N, Evans K, Lipsitz J, Olin J, Pearson J, Kalali A. The GRID-HAMD: standardization of the Hamilton Depression Rating Scale. Int Clin Psychopharmacol. 2008;23:120–129. doi: 10.1097/YIC.0b013e3282f948f5

47. Nakonezny PA, Carmody TJ, Morris DW, Kurian BT, Trivedi MH. Psychometric evaluation of the Snaith-Hamilton pleasure scale in adult outpatients with major depressive disorder. Int Clin Psychopharmacol. 2010;25:328–333. doi: 10.1097/YIC.0b013e32833eb5ee

48. Keser Z, Ikramuddin S, Shekhar S, Feng W. Neuromodulation for Post-Stroke Motor Recovery: a Narrative Review of Invasive and Non11Invasive Tools. Curr Neurol Neurosci Rep. 2023;23:893–906. doi: 10.1007/s11910-023-01319-6

49. Cole EJ, Phillips AL, Bentzley BS, Stimpson KH, Nejad R, Barmak F, Veerapal C, Khan N, Cherian K, Felber E, et al. Stanford Neuromodulation Therapy (SNT): A Double-Blind Randomized Controlled Trial. Am J Psychiatry. 2022;179:132–141. doi: 10.1176/appi.ajp.2021.20101429

